# Implementation of the 7-1-7 Framework through an Early Action Review of a Cholera Outbreak in Migori County, Kenya

**DOI:** 10.1101/2025.07.29.25332335

**Authors:** Naomi Roosevelt, Clement Ayungo Odero, Tom Odhong, James Oduor, Kenneth Okumu, Peter Okello, Boniface Olalo, Evance Ogola, Allan Munema, Francis Nga’nga, Catherine Kiama, Michelle Wangui, Esther Bundi, Nicholas Kurgat, Wycliffe Matini, Caleb Chemirir, Paul Olale, Naomi Ngaruiya, Marselah Chrisnah, Audrey Nasirumbi, John Ogange, Ahmed Fidhow, Daniel Langat, Grace Ikahu, Mark Nanyingi

**Author notes:** These authors contributed equally to this work (Naomi Roosevelt, Mark Nanyingi).

## Abstract

**Background:** Cholera remains a persistent public health threat in Kenya, exacerbated by inadequate water and sanitation infrastructure, cross-border mobility, and under-resourced health systems. On 12 February 2025, Migori County reported a cholera outbreak, prompting an Early Action Review (EAR) guided by the 7-1-7 framework to evaluate timeliness of detection, notification, and response.

**Methods:** We conducted a qualitative EAR involving review of outbreak timelines, epidemiological data, and facilitated focus group discussions with 40 stakeholders from county, national, and partner institutions. The outbreak chronology was benchmarked against the 7-1-7 metric: detection within 7 days, notification within 1 day, and response initiation within 7 days. Thematic analysis of six response pillars was combined with root cause analysis to identify bottlenecks and enabling factors.

**Findings:** The outbreak lasted 58 days (10 February–8 April 2025), resulting in 53 suspected cases, 16 confirmed, and one death (CFR 1.8%). All 7-1-7 performance targets were met: detection (2 days), notification (<1 day), and response (3 days). Key enablers included rapid diagnostic tests (RDTs), functional event-based surveillance, activated emergency operations centers, and buffer stocks of WASH and medical supplies. However, delayed culture confirmation and reliance on external partners for emergency supplies were noted as critical gaps.

**Interpretation:** Migori County’s application of the 7-1-7 framework through an EAR enabled a timely and coordinated cholera response. Institutionalizing EARs and strengthening laboratory and logistics capacity are essential to enhance epidemic preparedness and response in decentralized health systems.

## Introduction

Cholera, an acute diarrhoeal illness caused by toxigenic Vibrio cholerae serogroups O1 and O139, manifests with profuse watery diarrhea, vomiting, and muscle cramps within 2 hours –5 days of exposure. If untreated, it can cause severe dehydration and death within hours but with prompt recommended rehydration, the case-fatality rates fall below 1%(1). Globally, cholera remains endemic in many low- and middle-income countries, with an estimated 3–5 million cases and up to 143,000 deaths annually (1,2). From January to March 2025, WHO reported over 116,000 cases and nearly 1,500 deaths across 25 countries, predominantly in the African Region (66,689 cases, 1,336 deaths) and in the Eastern Mediterranean Region [3]. High-risk settings include rural, flood-affected, conflict and displaced-person camps with inadequate water, sanitation, and hygiene (WASH) infrastructure (3).

*V. cholerae* is transmitted via the faecal–oral route, primarily through ingestion of contaminated water or food. Shellfish and plankton are also known reservoirs; heavy rainfall, flooding, and breakdown of sanitation may disperse bacteria into human water sources (4). Transmission is amplified in overcrowded urban slums and camps; environmental and socio-economic drivers including climate variability further influence outbreak dynamics (5). In Africa, cholera epidemiology is marked by recurring outbreaks, often triggered by environmental shocks. A recent analysis noted an alarming resurgence in southern -eastern Africa, risking overshoot of 2021’s burden; only Kenya, Ethiopia, and Zambia were deemed on track for WHO’s 2030 elimination goal (6). Kenya has faced large-scale cholera outbreaks since 2014, including over 10,500 cases in 2015, over 5,200 in 2019, and around 12,100 by late 2023, with case-fatality rates averaging at 1.7% (7). Cholera cycles recur every 5–7 years, lasting 2–3 years, often tied to WASH deficits, flooding, food hygiene lapses, and overpopulation (8). Kenya’s current control strategy involves hotspot mapping, environmental surveillance, WASH improvements, oral cholera vaccination (OCV) campaigns, and multi-sectoral coordination (9,10). Prevention centers focuses on ensuring clean water, safe sanitation, hygiene education, and vaccination. Three WHO-prequalified OCVs Dukoral, Shanchol, and Euvichol show robust protection, particularly in endemic hotspots (4). WHO’s “Ending Cholera: A Global Roadmap to 2030” emphasizes rapid detection and response, WASH enhancements, and vaccine use in high-risk settings (1). However, persistent under-resourcing, weak infrastructure, and climate vulnerabilities continue to challenge progress.

On 12^th^ February 2025, Migori County reported a cholera index case, cases were detected in Kuria East, Kuria West, Suna East, and Suna West. Between 12th February and 2nd April 2025, the county recorded a total of 53 suspected cholera cases, and 1 death. An Early Action Review (EAR) was conducted to to document lessons learned and improve future response readiness. The EAR used the 7-1-7 framework, a global metric for evaluating the timeliness of outbreak response to epidemics (11).

## Methods

### Setting and outbreak overview

Migori County is located in southwestern Kenya, along the border with Tanzania, and is part of the former Nyanza Province. It shares borders with Homa Bay County to the north, Kisii County and Narok County to the east, Lake Victoria to the west, and Mara Region of Tanzania to the south. The county is a key cross-border trade and migration corridor, with several formal and informal border points, including the Isebania One Stop Border Post, which increases its vulnerability to cross-border disease transmission. The county is administratively divided into eight sub-counties, including Kuria East, Kuria West, Suna East, Suna West, Nyatike, Rongo, Awendo, and Uriri. It has a population of over 1.2 million people, with diverse rural and peri-urban settlements, and many areas with limited access to improved water and sanitation infrastructure (Figure 1).

**Figure 1:**
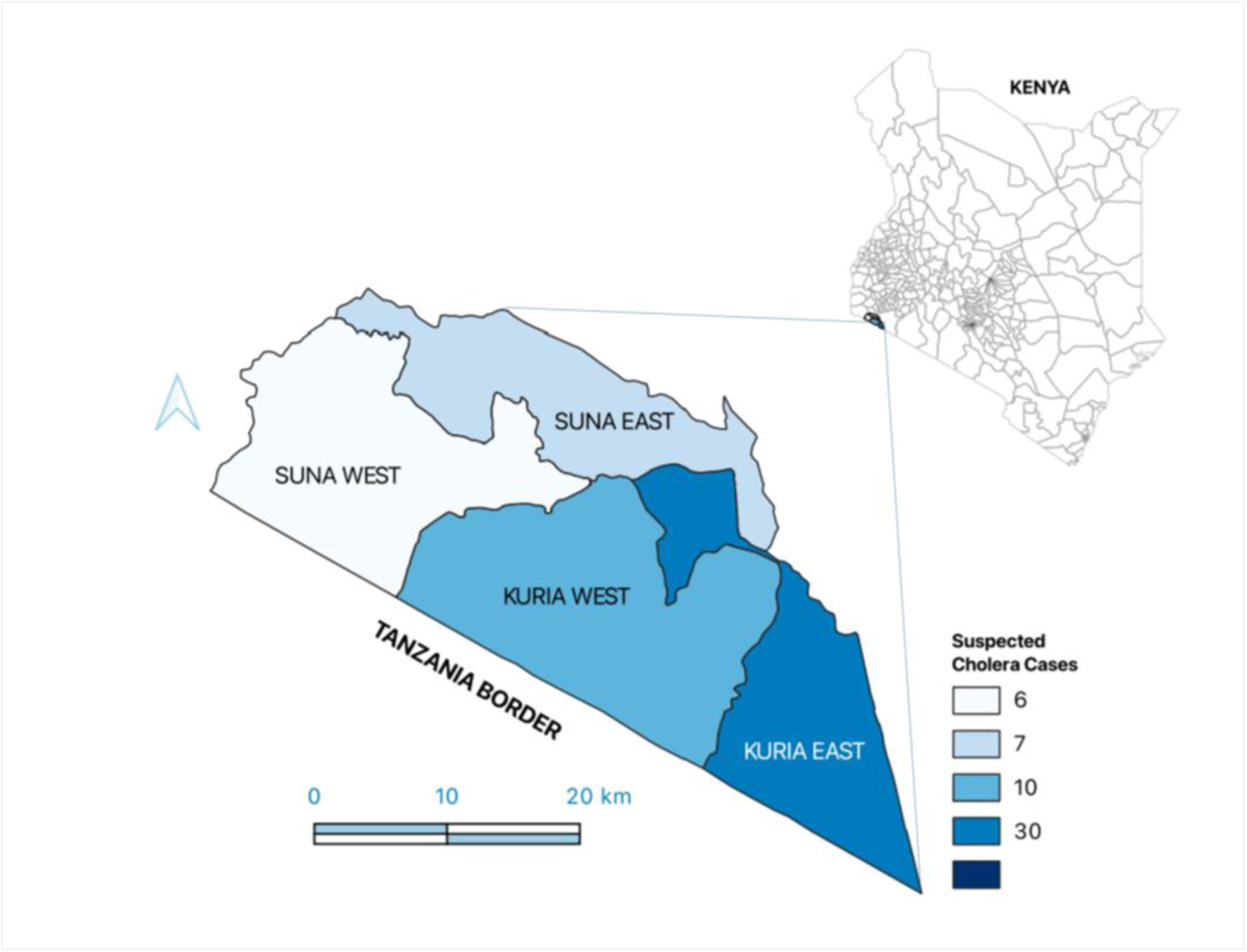
Spatial distribution of suspected cholera cases in Migori County, Kenya (Feb– Apr 2025) The map highlights the four most affected sub-counties Kuria East, Kuria West, Suna East, and Suna West using a graduated choropleth scale to reflect case intensity. The map also underscores the cross-border transmission risk posed by proximity to the Tanzania border, particularly in Kuria East and Kuria West. This necessitated enhanced community event-based surveillance (CEBS) and cross-border coordination with health authorities on both sides.

### EAR framework and design

The EAR followed structured guidance recommended by the World Health Organization (WHO) and Resolve to Save Lives (RTSL) for reviewing outbreak responses (12–14). The process assessed system performance across three critical milestones: Detection of the outbreak within 7 days of symptom onset, Notification to relevant authorities within 1 day of detection and Response initiation within 7 days of notification. These 7-1-7 targets provide a global standard for measuring the timeliness of epidemic responses and have been adopted by over 30 countries to improve health system performance (15)

### Stepwise EAR process

The qualitative review was conducted through a five multi-stage process:

#### Planning

Four virtual planning sessions were convened with stakeholders from the County Department of Health, Ministry of Health, and implementing partners (Kenya Red Cross Society, Washington State University, US CDC, AFENET, WHO). An exercise management team of subject matter experts reviewed existing tools based on the WHO EAR guidance and adapted them to the Kenyan subnational context.

#### Data Collection and Chronology Mapping

During the workshop, the epidemiological data from the county situational reports (SITREP) were reviewed by a multidisciplinary team to construct a timeline of outbreak events, including case onset, detection, reporting, and public health actions.The epidemic curve was used to anchor the assessment against 7-1-7 milestones (16).

#### Focus Group Discussions (FGDs)

Four FGDs, each comprising at least 10 health experts in various cadres were constituted to for facilitated discussions during a two-day workshop (3^rd^-4^th^ April 2025). The participants included of 24 health officers from the Migori county Department of Health Services, One port health officer from Isebania One Stop Border Post (OSBP), Five officers from the national Ministry of Health and10 officers from partner organizations. The discussants were subject matter experts in epidemiology, surveillance, emergency preparedness and response, risk communication and community engagement, WASH, clinical care and IPC. The discussions were led by four facilitators and one note taker documented the entire process.

#### Thematic analysis of key response pillars

Discussions were structured around six pillars of outbreak response: Surveillance, Coordination, Risk Communication and Community Engagement (RCCE), Case Management and Infection Prevention and Control (IPC), Water, Sanitation and Hygiene (WASH) and Laboratory Systems (17)

#### Root cause analysis and synthesis

A facilitated root cause analysis was used to identify bottlenecks, enabling factors, and best practices across pillars. The findings were synthesized into actionable recommendations, categorized into immediate and long-term priorities.

## Results

On 12 February 2025, Migori County reported its index cholera case in Kuria East Sub-county, confirmed initially by a rapid diagnostic test (RDT) and later by bacterial culture. Over the course of the outbreak, which spanned 58 days (from 10 February to 8 April 2025), a total of 53 suspected cases were reported across four sub-counties Kuria East, Kuria West, Suna East, and Suna West. Kuria East recorded the highest burden, with an attack rate of 1.9%, while Kuria West, Suna East, and Suna West each reported lower but epidemiologically linked clusters. Geographically, case clustering was observed around Namba, Sakuri B, and Posta villages, which are associated with informal mining activity, poor sanitation, and high population density. The sub-counties affected were also close to transboundary trade routes, underscoring the importance of integrated cross-border surveillance and localized Case Area Targeted Interventions (CATI) in these hotspots (Figure 2).

**Figure 2:**
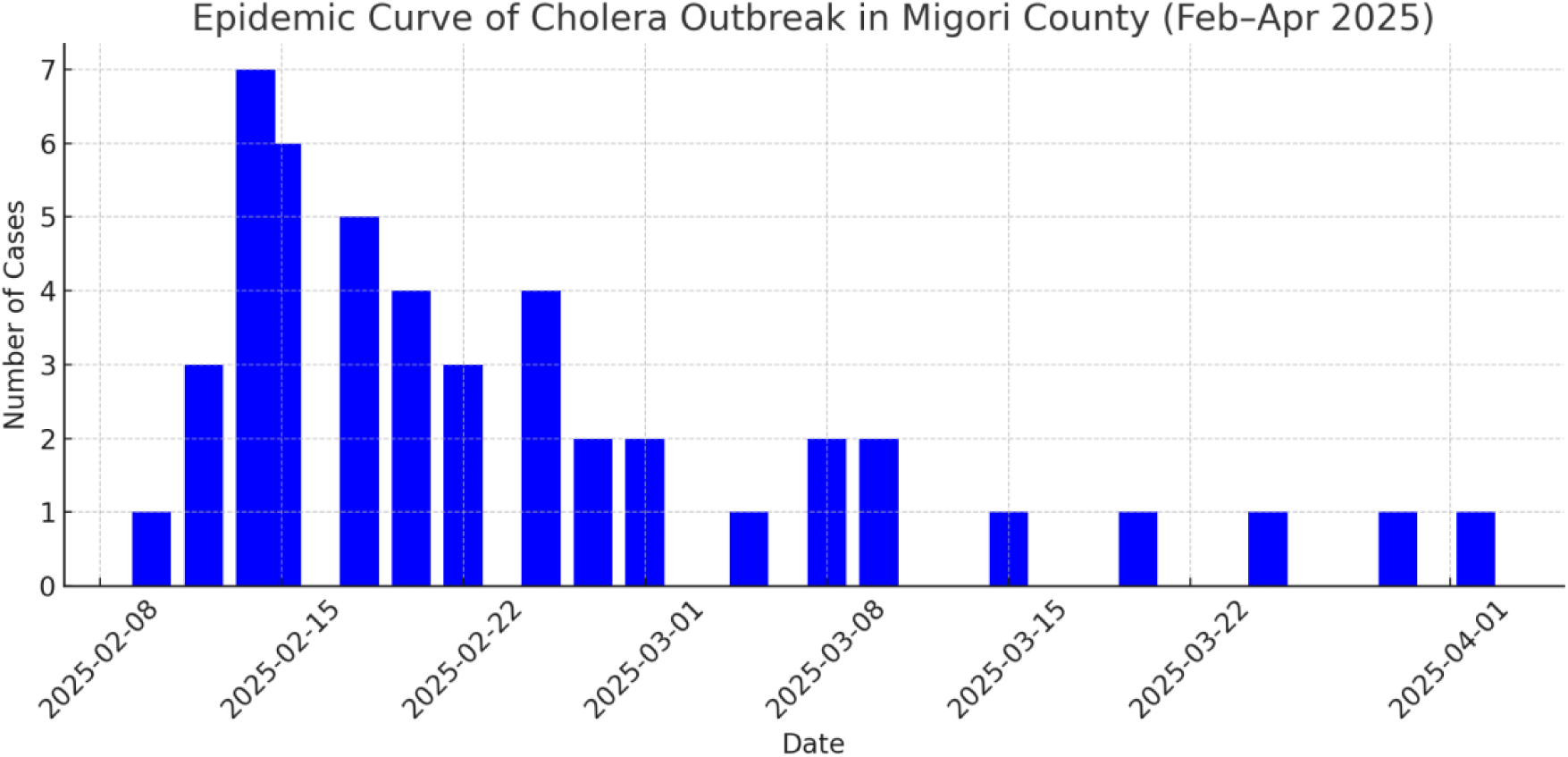
Epidemic curve of the cholera outbreak in Migori County, Kenya (February– April 2025), showing the temporal distribution of reported cases. Epidemic curve of the cholera outbreak in Migori County from 10 February to 8 April 2025. The curve illustrates the temporal distribution of 53 suspected cases. A sharp increase was observed around 21 February, followed by a gradual decline after early public health interventions including the deployment of rapid response teams and WASH activities.

Among the suspected cases, 43 tested positive using Rapid Diagnostic Tests (RDTs), while 16 cases were confirmed by culture for *Vibrio cholerae* O1, serotype Ogawa. The outbreak resulted in one death among the 37 probable cases, producing a case fatality rate (CFR) of 1.8%, which is slightly above the WHO-recommended threshold of <1% but significantly lower than in many regional outbreaks, suggesting relatively effective case management interventions. The epidemic curve shows that the outbreak reached its peak around 21 February 2025, approximately 11 days after the first symptoms were reported. This peak corresponds with a surge in suspected cases, after which the incidence began to decline steadily (Figure 3). This trend aligns temporally with the activation of the County Incident Management System (IMS) and the deployment of Rapid Response Teams (RRTs), which began on 15 February 2025.

**Figure 3:**
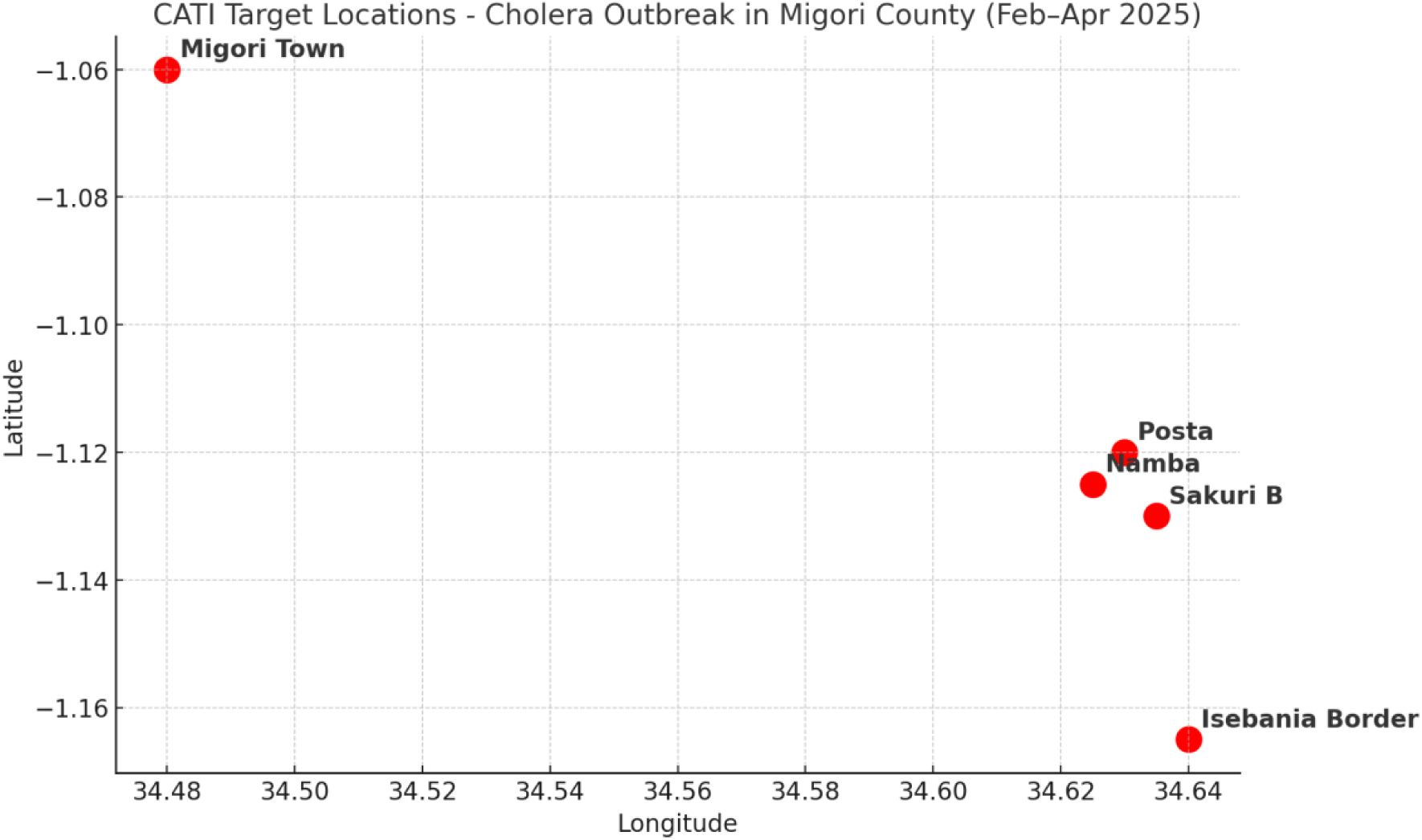
Case Area Targeted Interventions (CATI) map of affected villages in Migori County, Kenya. This spatial visualization informed the implementation of **Case Area Targeted Interventions (CATI)**, a focused outbreak control strategy that deploys rapid, localized measures around confirmed or probable cases. These interventions included:

- Household-level WASH kits distribution (chlorine, soap, water containers),
- Emergency latrine repairs and desludging,
- Door-to-door risk communication and health education,
- Active case finding and referral,
- Chlorination of local water sources. By identifying cholera hotspots, this CATI map enabled the Migori County Department of Health **a**nd partners to deploy limited resources more efficiently, breaking transmission chains early and limiting the geographic spread of the outbreak.

**Figure 4:**
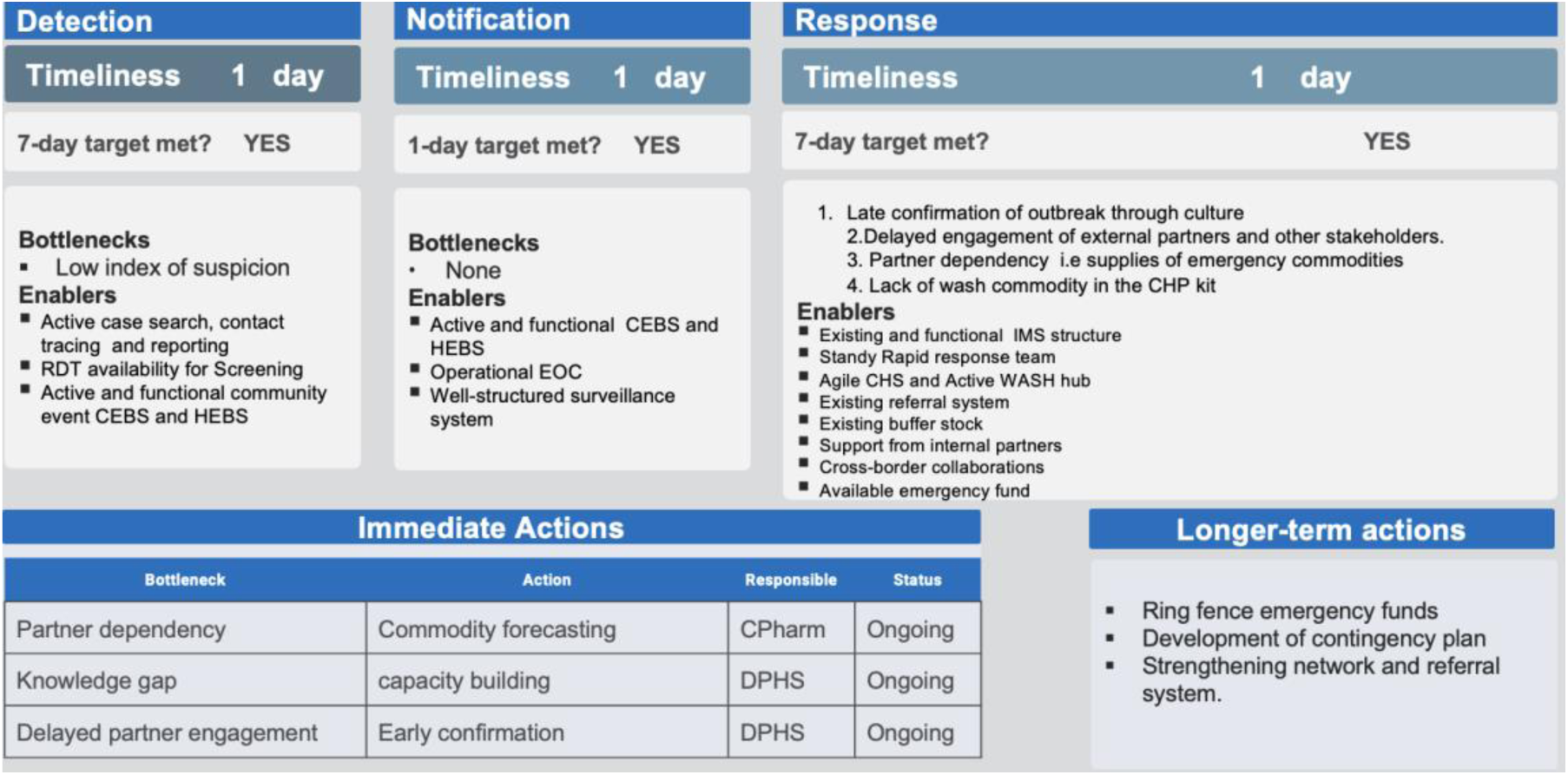
Summary of Migori County’s Early Action Review (EAR) of the February–April 2025 cholera outbreak using the 7-1-7 performance framework. The figure outlines the timeliness of detection, notification, and response, along with corresponding bottlenecks, enablers, and mapped corrective actions. All 7-1-7 targets were achieved, demonstrating system functionality under pressure.

### 7-1-7 performance

The 7-1-7 framework, which sets benchmarks for detecting a public health event within 7 days of onset, notifying relevant authorities within 1 day of detection, and initiating a response within 7 days of notification, was used to assess the timeliness of Migori County’s cholera outbreak response. The county met all three targets, demonstrating operational readiness in surveillance and response mechanisms as outlined below and in (**Error! Reference source not found.**):

### Detection (within 2 days of symptom onset)

The index case, a resident of Kuria East Sub-County, exhibited symptoms on 10^th^ February 2025 but succumbed the same day, a suspected cases that were in contact with the index case was identified through syndromic surveillance at a local health facility. The case was screened and confirmed presumptively using a Rapid Diagnostic Test (RDT) by 12^th^ February 2025. This early detection was facilitated by an active Community Event-Based Surveillance (CEBS) system, which enabled timely referral from the community to the health facility by the community health promoters (CHP) and the community health assistants (CHA). The use of decentralized RDTs and high index of suspicion among facility staff were key enablers in achieving this prompt detection.

### Notification (on the same day as detection)

Immediate notification protocols were followed on 12^th^ February 2025, with the facility reporting the suspected case to the County Emergency Operations Center (EOC) and onward to the national Public Health Emergency Operations Center (PHEOC). This was supported by an established Incident Reporting and Alert System (IRAS), which allows rapid submission of SPOTREP (Spot Reports) and other situational reports (SITREP) and updates. Timely notification ensured early decision-making and coordination of response efforts. The notification was aligned with Integrated Disease Surveillance and Response (IDSR) protocols.

### Response (initiated within 3 days of detection)

Full-scale response activities were triggered on 15^th^ February 2025, including deployment of the County Rapid Response Teams (RRT), distribution of emergency WASH and medical commodities to affected sub-counties, and the commencement of outbreak investigation. An initial rapid risk assessment was conducted, and Case Area Targeted Interventions (CATI) were implemented in hotspots such as Namba, Sakuri B, and Posta villages (Figure 2). The functional Incident Management System (IMS), availability of pre-positioned supplies, and access to emergency funds contributed to meeting the 7-day response benchmark.

**Table 1:**
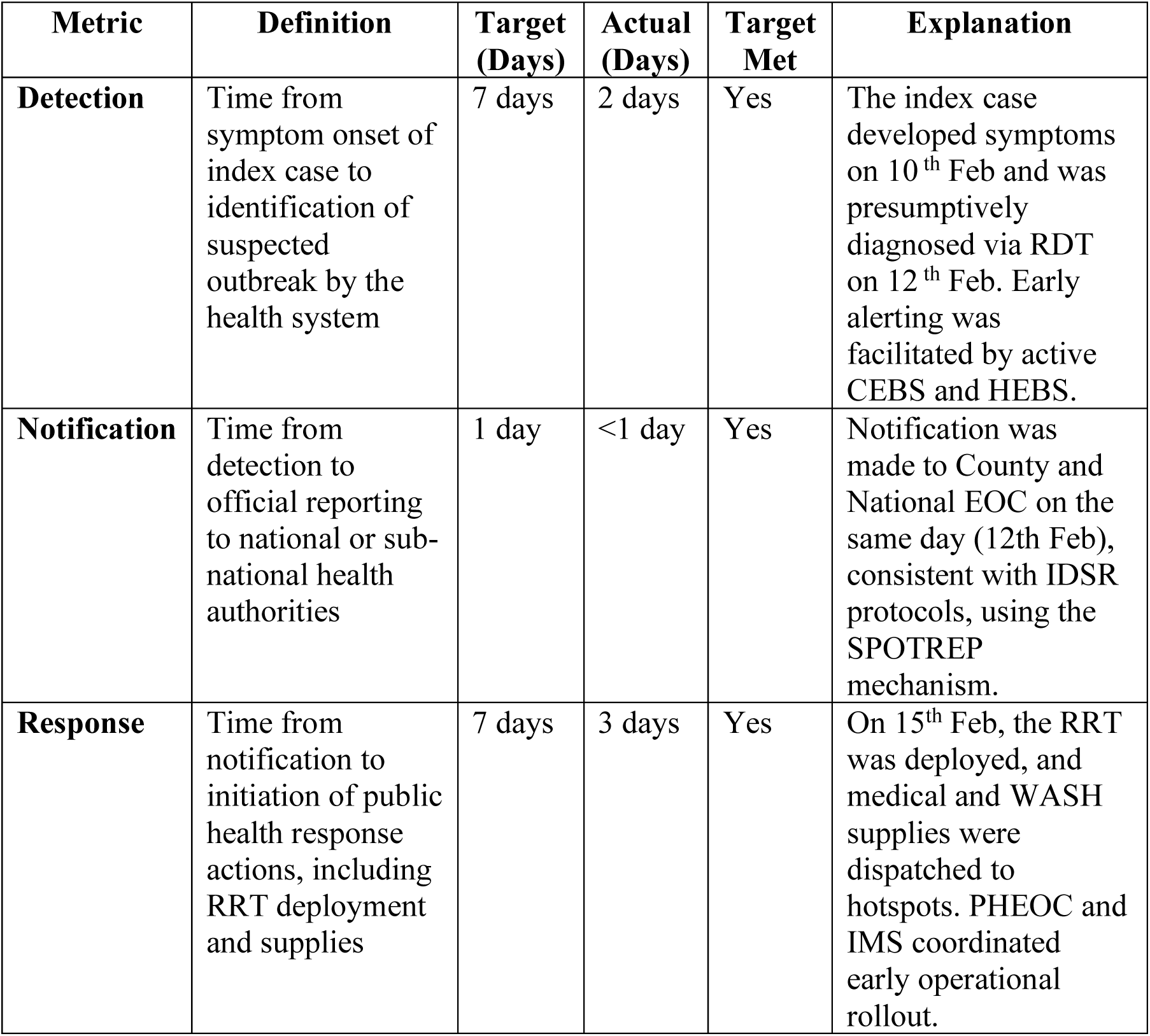
7-1-7 Performance Metrics.

### Best Practices

The EAR identified several best practices and enabling factors that contributed to the successful early response to the cholera outbreak in Migori County. These practices can serve as reference models for similar settings, they include:

#### Availability and Use of Rapid Diagnostic Tests (RDTs)

The presence of RDT kits at the health facility level allowed for prompt screening and presumptive diagnosis of suspected cholera cases. This accelerated initial public health interventions while awaiting culture confirmation. The integration of RDT use into standard triage procedures proved critical in minimizing delays in case detection and isolation.

#### Active Community and Hospital Event-Based Surveillance (CEBS/HEBS)

A functional surveillance architecture combining the detection and reporting of signals by CHPs and CHAs and facility-level alerts enabled the early flagging of the index case. CHPs actively engaged in detecting and reporting suspected cases, while healthcare facilities maintained vigilance through syndromic surveillance. This dual surveillance system enhanced the sensitivity of early detection mechanisms.

#### Rapid Mobilization of Response Teams

The pre-designated Rapid Response Teams (RRTs) were promptly deployed following case confirmation. This quick activation was supported by the existing Incident Management System (IMS) and clear response protocols, allowing for immediate outbreak investigation, contact tracing, and risk assessments.

#### Operational Public Health Emergency Operations Center (PHEOC)

Migori County’s EOC functioned as a coordination hub, facilitating multi-sectoral engagement and streamlined communication between county EOC, national PHEOC, and partner stakeholders. The County EOC maintained real-time incident tracking and ensured harmonized task allocation across all response pillars.

#### Pre-positioned WASH and Medical Commodities

The presence of buffer stocks for essential supplies such as chlorine, soap, water containers, IV fluids, and antibiotics enabled rapid deployment of resources to affected areas. These stocks were available at sub-county depots and were dispatched based on early assessments, reducing dependence on external procurement in the first phase.

#### Cross-Border and Multi-Sectoral Collaboration

Engagement with neighboring Tanzania through the Isebania One Stop Border Post (OSBP) facilitated timely information sharing and joint response strategies through population mobility monitoring. Additionally, the involvement of education, water, disaster management, and local governance structures bolstered the comprehensiveness of response measures.

#### Targeted Risk Communication and Community Engagement (RCCE)

The RCCE strategy utilized trusted community structures, including chiefs, CHPs, KRCS trained volunteers and local FM radio stations, to deliver consistent health messages. These interventions were responsive to community concerns, culturally tailored, and effective in promoting uptake of preventive practices such as handwashing, safe water use, and early care-seeking

### Bottlenecks and Enablers

The EAR revealed several operational and systemic challenges that impacted the optimal performance of the cholera outbreak response in Migori County. They are summarized below, while **Error! Reference source not found.** summarizes the respective enablers:

#### Delayed Culture Confirmation

Although RDTs enabled prompt initial detection, there was a notable delay in confirmatory testing via bacterial culture. Culture confirmation took up to 9 days post-sample collection, exceeding ideal timelines for early outbreak classification and risk communication. This delay was attributed to limited laboratory capacity, constrained logistics for sample transport, and backlog at reference laboratories.

#### Inadequate Contingency Stock for Laboratory Commodities

The county experienced shortages in critical laboratory supplies including culture media, reagents, and transport materials during the early days of the outbreak. These gaps hampered timely processing and verification of samples and highlighted a lack of pre-positioned contingency stock, which is essential for maintaining uninterrupted diagnostic services during emergencies.

#### Knowledge Gaps among Frontline Clinicians

Some healthcare providers demonstrated a low index of suspicion for cholera during the initial phase, leading to missed opportunities for early case identification and triage. This gap was linked to limited prior training on epidemic prone and IDSR priority diseases, especially in sub-counties that had not experienced previous cholera outbreaks. The absence of targeted refresher courses further exacerbated this knowledge gap.

### Overdependency on partners for emergency supplies

The response was heavily reliant on external partners for key emergency commodities such as WASH kits, IV fluids, and Personal Protective Equipment (PPE). While these partners played a vital role, the dependency created logistical delays and limited county ownership and flexibility in response actions, particularly in the initial 72 hours.

### Lack of Standardized Job Aids, Guidelines, and SOPs

Response teams, particularly at the sub-county and facility levels, lacked access to updated cholera-specific protocols and job aids. This affected consistency in case management, sample collection procedures, reporting, and risk communication. The absence of operational guidance materials contributed to variability in response quality and reduced adherence to national standards

### 3.2 Response action completion

The EAR assessed the timeliness and quality of core public health response actions implemented during the initial phase of the cholera outbreak. The review mapped each action against the response timeline to determine whether it was completed within the 7-day response window, in accordance with the 7-1-7 framework (Table 2). Of the seven key response pillars assessed, six met the expected threshold for both timeliness and operational coverage. These included:

- Outbreak investigation and rapid risk assessment, which were initiated within 72 hours of case detection,
- Case management and infection prevention and control (IPC), facilitated through the activation of temporary isolation wards and the deployment of trained clinicians,
- Medical countermeasures (MCMs) and WASH interventions, enabled by the availability of pre-positioned emergency stocks and buffer supplies at the county warehouse,
- Risk communication and community engagement (RCCE) activities, which leveraged community health promoters, local FM stations, and chiefs’ barazas to deliver hygiene and care-seeking messages,
- Coordination mechanisms, particularly through the activation of the County EOC and Incident Management System (IMS), which enabled real-time decision-making and partner mobilization demonstrating the benefits of proactive emergency governance.

**Table 2:** Early Response Action Completion.

| **Response Action** | **Description** | **Target Met** | **% Met Target** |
| --- | --- | --- | --- |
| **Initiate Investigation** | Immediate deployment of surveillance and field epidemiology teams to conduct outbreak investigation, case verification, and contact listing. | Yes | 100% |
| **Epidemiological Characterization & Risk Assessment** | Profiling of cases, generation of an epidemic curve, spatial clustering analysis, and rapid risk assessment to guide interventions. | Yes | 100% |
| **Laboratory Confirmation** | Culture confirmation of *Vibrio cholerae* O1/Ogawa at a certified laboratory. Despite delays, sufficient samples were confirmed to validate outbreak. | Partially | 71% |
| **Case Management and IPC** | Setup of temporary isolation units, deployment of trained staff, and availability of IV fluids, ORS, and antibiotics at designated facilities. | Yes | 100% |
| **Medical Countermeasures (MCMs) / PHSMs** | Timely provision of emergency medical supplies, activation of public health and social measures (e.g., mass handwashing stations, movement advisories). | Yes | 100% |
| **Risk Communication and Community Engagement (RCCE)** | Community sensitization using local media, chiefs, and CHPs; delivery of cholera prevention messages in hotspot areas. | Yes | 100% |
| **Coordination Mechanism** | Activation of the County Public Health Emergency Operations Center (PHEOC), Incident Management System (IMS), and joint partner coordination. | Yes | 100% |

## Discussion

Cholera continues to pose a significant threat to public health security across sub-Saharan Africa, where it is fueled by a convergence of environmental, socio-political, and structural vulnerabilities. Recent analyses have revealed that persistent transmission foci remain concentrated in regions with chronic WASH deficits, population displacement, and fragile health systems.The progress toward the 2030 elimination roadmap will require addressing the main drivers including weak governance structures, poor integration of environmental data, and urban informal settlements. These insights underscore the need for localized outbreak reviews such as this Early Action Review (EAR) from Migori, to unpack operational realities and refine subnational preparedness models (18,19)

This EAR demonstrates the feasibility and effectiveness of implementing the 7-1-7 performance framework to assess and improve early response to a cholera outbreak in a subnational setting. Migori County met all three performance targets-detection within 7 days, notification within 1 day, and response within 7 days underscoring the operational readiness of its surveillance, coordination, and response mechanisms. These results are consistent with findings from other countries that have applied the 7-1-7 framework to accelerate public health response cycles and identify systems-level gaps in real time (11,12)

A key factor in the success of Migori’s response was the timely detection of the index case, facilitated by active Community and Hospital Event-Based Surveillance (CEBS/HEBS) systems. The use of Rapid Diagnostic Tests (RDTs) at peripheral health facilities enabled presumptive diagnosis within 48 hours, significantly contributing to early outbreak recognition and containment. The concurrent activation of the County Emergency Operations Center (EOC), deployment of Rapid Response Teams (RRTs), and release of buffer stocks for WASH and medical supplies reflect a coordinated, multi-sectoral approach that aligned with international best practices of outbreak containment (5,20).

However, the EAR revealed challenges that mirror those observed in similar decentralized settings(6). Most notably, the delay in laboratory confirmation taking up to nine days highlighted gaps in contingency planning for diagnostic capacity and logistics. Similar delays have been reported in Uganda and Nigeria, where limited availability of culture media and constrained referral systems reduced laboratory efficiency during cholera and Lassa fever outbreaks. Addressing these bottlenecks requires targeted investment in laboratory preparedness, including pre-positioned supplies and trained personnel at the county level(11).

The Migori County EAR was grounded in a structured review of 7-1-7 performance metrics, allowing for evaluation against both national guidelines and global performance expectations. Notably, this approach aligns with emerging evidence on the effectiveness of early warning systems and decentralized preparedness reviews in the Kenyan context. Previous studies have emphasized that counties with functional early detection systems particularly those leveraging Rapid Diagnostic Tests and local EOCs achieved significantly faster containment outcomes during recent cholera surges(21).

The implementation of the 7-1-7 framework also offered valuable insights into institutional readiness and response accountability. WHO and Resolve to Save Lives have promoted the 7-1-7 framework as a global organizing principle for epidemic preparedness, now adopted by more than 30 countries (11,13).Migori’s experience reinforces its utility at the subnational level, particularly when integrated with routine After-Action Reviews (AARs) and Early Action Reviews (EARs)(15). An operational case study from Garissa County reinforces the utility of structured cross-border surveillance, pre-positioned buffer stocks, and community-centric coordination practices echoed in the Migori response(22). At a continental scale, only 61% of African countries have implemented cholera preparedness plans, with Kenya among a handful integrating the 7-1-7 model at both national and subnational levels(23). The Migori EAR thus contributes essential evidence of how localized frameworks can operationalize global targets under real-world constraints ((21–23)).

The use of EARs as a learning tool also proved beneficial. Though conducted 52 days into the outbreak, the exercise provided a structured platform for joint problem-solving, multi-agency reflection, and prioritization of corrective actions. It also contributed to a culture of continuous improvement within the health system, strengthening local leadership and adaptive planning. A recent study in Uganda provides a broader systems-level perspective on institutionalizing the 7-1-7 framework across multiple outbreaks, offering an instructive benchmark for subnational preparedness reviews like Migori’s. Notably, both countries reported similar structural barriers limited diagnostics, resource constraints, and geographic remoteness yet Uganda’s integrated EOCs and regular simulation exercises allowed for more consistent adherence to 7-1-7 targets anchoring on value of institutional scaffolding that serve as a model for scaling EARs in Kenya (24).

## Recommendations

The EAR outlined a suite of strategic recommendations to improve cholera outbreak preparedness and response grounded in both observed bottlenecks and successful interventions during the 2025 response. They include both immediate and long-term actions as outline below and summarised in (**Error! Reference source not found.**)

### Immediate Actions

#### Health workforce training

Targeted in-service training, mentorship, and continuous professional development for frontline health workers are urgently needed. These efforts should focus on clinical case recognition, triage, sample collection, use of diagnostic tools, and cholera-specific management protocols. Orientation sessions on the 7-1-7 framework and IDSR guidelines should be prioritized at the sub-county and facility levels.

#### Timely laboratory culturing

Investments in peripheral laboratory capacity and personnel training are essential to reduce delays in culture confirmation. Strengthening referral pathways for specimens, ensuring cold-chain integrity, and equipping facilities with appropriate culture media and reagents can significantly improve diagnostic turnaround time.

#### Restocking and logistics optimization

A dynamic supply chain mechanism should be instituted to facilitate continuous assessment, forecasting, and redistribution of emergency commodities including RDTs, IV fluids, ORS, chlorine tablets, and PPE. Real-time stock visibility at sub-county stores can be achieved through digital tools or regular reporting.

#### Distribution of SOPs and Job Aids

Standard operating procedures (SOPs), algorithms, and visual job aids for case management, sample handling, surveillance reporting, and community engagement should be disseminated to all health facilities. These materials must be context-specific, user-friendly, and harmonized with national cholera response protocols.

### Long-Term Actions

#### Ring-Fencing emergency funds

The County Government should institutionalize the allocation of emergency health response funds within the County Integrated Development Plan (CIDP) and annual budgets. Clear disbursement protocols and accountability mechanisms are necessary to ensure swift access to funds during future outbreaks.

#### Development of a Cholera contingency plan

A county multi-sectoral cholera contingency plan, aligned with the National Cholera Multi-sectoral Elimination Plan (NCMEP), should be developed, validated, and disseminated. The plan should include clear roles for stakeholders, simulation exercises, early warning triggers, and thresholds for activating response mechanisms.

#### Strengthening referral systems

Establishing efficient two-way referral and feedback systems between community units, peripheral health facilities, and referral hospitals will enhance case management and surveillance linkages. This includes improving transport, communication, and specimen tracking systems.

#### Enhancing Water infrastructure and quality monitoring

Long-term cholera prevention requires sustained investments in water infrastructure, including borehole rehabilitation, water treatment stations, and piped water networks in underserved communities. Continuous water quality testing, chlorination programs, and inter-sectoral WASH coordination are essential to address structural drivers of transmission.

Overall, in order to ustain early detection and coordinated response during future outbreaks, there is a need of institutionalizing Early Action Reviews and embedding the 7-1-7 metric within county-level epidemic preparedness plans. Continuous workforce capacity-building is pivotal, that targeted health education interventions significantly enhance frontline preparedness, particularly when delivered through continuous professional development and simulation-based exercises (SIMEX).

Migori County would benefit from investing in standardized RCCE protocols, including culturally tailored materials and trusted community networks, which have been shown to improve risk perception and hygiene behaviors.There is a need for advocacy to pre-allocate emergency funds, routine simulation exercises, and strengthened national-local coordination to mainstream rapid response systems to solidify Migori’s emerging leadership in decentralized outbreak control(25)

## Conclusions

Migori County’s successful implementation of the 7-1-7 framework through an Early Action Review offers an actionable model for outbreak preparedness and response in decentralized health systems. The county’s ability to detect, notify, and respond to a cholera outbreak within internationally accepted timeframes illustrates the potential of coordinated subnational systems to achieve global health security goals. While areas for improvement remain particularly in laboratory diagnostics the county’s performance underlines the importance of investing in surveillance architecture, emergency logistics, and multi-sectoral coordination. Institutionalizing EARs and adopting 7-1-7 metrics at national and subnational levels can catalyze rapid performance improvement and position counties like Migori at the forefront of epidemic intelligence and action.

## Data Availability

All data produced in the present study are available upon reasonable request to the authors

## Acknowledgements

We acknowledge the Migori County Department of Health, National Ministry of Health, University of Nairobi Washington State University, WHO, and all supporting partners for their technical support. Specifically the Kenya Red Cross for providing both logistical and technical support to the workshop. Special appreciation to the following individuals for their active participation and invaluable inputs during the exercise, Migori County: *Margaret Malowa, Dylphina Obino, Benard Onyango, James Oguk, Beffy Vill and Nyamohanga David*.

## Data Availability Statement

All relevant data supporting the findings of this study are available from the Migori County Department of Health and the Ministry of Health, Kenya. Aggregated anonymized outbreak data can be provided upon reasonable request.

## Ethical Review Statement

The ethics oversight for this study was provided by the Kenyatta National Hospital – University of Nairobi Ethics and Research Committee (KNH-UoN ERC), Nairobi, Kenya. The KNH-UoN ERC waived ethical approval, as the study was part of a Ministry of Health-led Early Action Review (EAR) conducted in response to a public health emergency. According to Kenyan legislation and established procedures, such outbreak response evaluations that do not involve invasive procedures or identifiable personal data are exempt from ethical review.

## Author Contributions

NR, CAO, TO, JO, KO, PO, BO, EO, AM and led outbreak investigation and coordination. MN, FN and PO provided technical support in evaluation design and implementation. CK, MW, EB, NK, PO, NN, CC, MC, AN, JO, and WM contributed to the Early Action Review facilitation. NR,TO, JO, AM and MN assisted in data collation and analysis. AF, DL, and GI contributed to interpretation and manuscript refinement. MN conceptualized the exercise and is the corresponding author and coordinated manuscript drafting. All authors reviewed and approved the final manuscript.

